# INTESTINAL PARASITES AND ANAEMIA AMONGST ADULT PATIENTS ATTENDING A TERTIARY HOSPITAL IN CALABAR, NIGERIA

**DOI:** 10.1101/2025.06.20.25329997

**Authors:** E. O. Effanga, E. E. Imalele, O. E. Okon

**Affiliations:** Department of Zoology and Environmental Biology, Faculty of Biological Sciences, University of Calabar, Calabar. Nigeria

**Keywords:** Anaemia, Intestinal, Parasites, Hospital, Calabar

## Abstract

A study was carried out to determine the prevalence of intestinal parasitic infestations and anaemia among adult outpatients (aged 21-50 years) attending the University of Calabar Teaching Hospital, Calabar, Cross River State, Nigeria. Anaemic status was determined by haematocrit while parasitic infestation was determined by floatation and direct smear techniques. 42% of the anaemic patients were positive for intestinal parasites while 4% of the non-anaemic patients were positive for intestinal parasites. This was very highly significant (x^2^=40.768, p<0.00001). Hookworm was the commonest parasite infecting patients (14%), followed by *Entamoeba histolytica* (9%), *Ascaris lumbricoides* (5%), *Trichuris trichiuria* (3.5%) and *Balantidium coli* (1%). Infections with hookworm were of moderate to high intensity. 26.6% males were infected as opposed to 18.7% females, though this was not statistically significant (X^2^=1.758 p=0.185). Polyparasitism was common in males than females and all cases of polyparasitism were anaemic. The age group 31-40 years had the highest infection rate (24.3%). The study showed a relationship between parasitic infestation and alteration of haematological indices. The results also highlighted that parasite control programs targeted at school children should be extended to people of all age groups. The anaemic conditions were worsened by parasite infestations.

## INTRODUCTION

Anaemia is the second most common cause of disability in the world [1], and is reflected in several of the global Millennium Development Goals [2]. More than two billion individuals (about one third of the world’s population) are anaemic due to several causes [3]. The main causes of anaemia are dietary iron deficiency, infectious diseases such as malaria, hookworm infections and schistosomiasis; deficiencies of other key micronutrient including folate, vitamin B_12_ and vitamin A; or inherited conditions that affect red blood cells (RBCs), such as sickle cell anaemia and thalassemia [4]. [5] reported that the etiology of anaemia in developing countries is multifactorial since it is seldom present in isolation but coexists with a number of other conditions like deficiencies of micronutrients (e.g., iron, folate, vitamin B_12_), haemoglobinopathies (thalassemia, cell disease), infection and infestation (e.g. malaria, hookworm, trichuriasis, ascariasis, schistosomiasis, HIV, tuberculosis, etc.), and cancer.

Globally the most significant contributor to the onset of anaemia is iron deficiency. Given the role of iron in oxygen transport and the low levels of available iron in the diets of a large proportion of the global population, iron deficiency anaemia (IDA) and anaemia are often used synonymously and the prevalence of anaemia has often been used as a proxy for iron deficiency anaemia [6]. Iron deficiency anaemia is in itself caused by a number of conditions which include insufficient dietary intake of iron, gastro-intestinal tract bleeding due to parasitic infections and malabsorption of nutrients in the intestines [3]. In addition, chronic blood loss as a result of menstruation, parasitic infections such as hookworm, ascariasis, acute and chronic infections including malaria, tuberculosis and HIV significantly lower blood haemoglobin concentration [6]. The health consequences associated with anaemia are most pronounced in children and women of reproductive age.

The presence of anaemia due to intestinal parasites has always been attributed to children and infants who seem less capable of avoiding the infectious agents, or because of increase in iron needs. Only few studies have been carried out to investigate the interplay of intestinal parasites and anaemia among the adult populace. This study is therefore aimed at determining the prevalence of intestinal parasites and evaluate the relationship between anaemia and parasitic infection among adult patients (21 - 50 years) attending the University of Calabar Teaching Hospital, Calabar.

## MATERIALS AND METHODS

### Study area

Calabar is the capital city of Cross River State in the coastal south-eastern region of Nigeria. It has an area of 406km^2^ and a population of 371, 022 at the 2006 census [7]. The city lies between latitude 4^0^ 57’’N and longitude 8^0^ 19’’E. It is divided into Calabar Municipal and Calabar South Local Government Areas. The study was carried out at the University of Calabar Teaching Hospital, Calabar was established in 1979 to serve as a tertiary Health Institution that will render clinical services at a level that meets the requirement for training medical students of the University of Calabar.

### Ethical clearance

The overall protocol of the study was approved by the Ethical Research Committee 0f the University of Calabar Teaching Hospital (U.C.T.H), Calabar. An informed consent form was signed by all the subjects included in the study.

### Subject recruitment

The study was open to participants aged 20-50 years. Subjects were recruited from the Out Patients Department (OPD) of the University of Calabar Teaching Hospital with the assistance of the medical staff. Interested participants were interviewed for compatibility, required to sign a consent form after which, their blood samples were collected for analysis by the departmental laboratory scientist. Specimen bottles were given to these patients so as to obtain their stool the following day for analysis. Based on their haematology result, the patients were divided into two groups - those with anaemia and those without anaemia (control). A total of 200 subjects (100 anaemic and 100 controls) were used for the study. The anaemic group had packed cell volume (PCV) values less than 39% for males and less than 33% for females according to WHO threshold for anaemia.

### Sample collection

#### Blood

Venous blood was obtained from each patient. The site for the collection of blood (the arm) was cleaned with 75% alcohol and allowed to dry. This procedure sterilises the skin and promotes a free flow of blood. A dry sterile syringe was used to draw blood from a suitable vein. The needle of the syringe was then removed and the blood slowly ejected into specimen tubes containing Ethylenediaminetetraacetic acid (EDTA) as anticoagulant. Adequate mixture of the blood and anticoagulant was achieved by repeated slow inversion.

Samples were obtained from males and females of ages 21 to 50 years over a period of eight months (July, 2009 - February, 2010).

#### Stool

Specimen bottles (with tight fitted caps) were given to patients with two applicator sticks to provide stool samples on their return to the hospital the following day.

#### Analysis of blood sample

Well-mixed anticoagulated blood was drawn into two micro-haematocrit tubes by capillary action. The tube was about ¾ filled. One end of each tube was sealed with clay material at 90^0^ angle. The sealed capillary tubes were then placed in a microhaematocrit centrifuge with the sealed ends pointing outside of the centrifuge. The duplicate sample was placed on the opposite end in order to balance the centrifuge. The lid was placed and the samples were spun for 5 minutes. The centrifuge was allowed to stop on its own before removing the tubes. The haematocrit (Packed Cell Volume) was measured with the haematocrit reader from the volume of erythrocytes as a percentage of the total sample volume in the capillary tubes [8][9]

### Stool analysis

#### Macroscopic examination of stool

As soon as the specimen was received in the laboratory, its nature (degree of formation) was noted with the following letters written on the container: F (semiformed), S (soft), L (loose), or W (watery). M was written if mucus was present and B if blood was present. This was done as a guide as to whether the trophozoite stage or the cyst stage of protozoa was likely to be present.

#### Wet mount

A drop of fresh physiological saline was placed on one end of a clean grease-free slide and a drop of iodine on the other end. Using a wire loop a small portion of the stool specimen was mixed with the saline. The wire loop was flamed before a similar amount of the same sample was mixed with the iodine. Each preparation was then covered with a coverslip. The prepared slide was mounted on the microscope and examined systematically for the presence of larvae, eggs, trophozoites, cysts and oocysts of parasites. The low-power (x10) objective was first used to focus on the mount. When organisms or suspicious material were seen, it was switched to the high-dry (x40) objective for detailed observation. The number of parasite-specific eggs per gram of stool (EPG) was used to define infections as low, moderate or high intensity in accordance with WHO-established intensity cut-off values for helminth infections [10][8][11][12].

#### Faecal floatation

Brine was used as the floatation solution. Four hundred grams (400g) of granular Sodium Chloride was dissolved in 1000ml water to produce a medium with a specific gravity of approximately 1.2. A wooden applicator stick was used to mix approximately 2grams of faecal sample in 10ml floatation solution. This was poured through a tea strainer into a beaker. It was then poured from the beaker into a 10ml centrifuge tube. More of the floatation solution was added to create a slight inverted meniscus. This was centrifuged at 1200rpm for 5minutes in a swinging-cup centrifuge. A coverslip was then placed on top and was left to stand for 10minutes after which the coverslip was removed, placed on a microscope slide and viewed under the microscope [8][13].

#### Determination of prevalence and intensity of parasites

All parasite eggs on the slide were counted and tallied for each type of egg identified. All counting and estimation assessed the number of parasitic elements (eggs, larvae, oocysts, cysts) per gram of faeces (EPG/LPG/OPG), and were based on the microscopic examination of faecal suspension from a known volume of a faecal sample. To determine the number of parasite eggs per gram of faeces (EPG) the formula below was used:

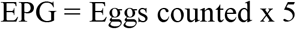

This is because 2gram sub-sample was diluted to 10ml. So a ratio of 1:5 weight/volume was created [13][14].

Furthermore, samples were subdivided into low, moderate, and high egg excretion intensities according to thresholds proposed by WHO; for *A. lumbricoides* these were 1– 4,999 EPG, 5,000–49,999 EPG, and 49,999 EPG; for *T. trichiura* these were 1–999 EPG, 1000–9,999 EPG, and >9,999 EPG; and for hookworm these were 1– 1,999 EPG, 2,000–3,999 EPG, and >3,999 EPG, respectively. Very high worm load for hookworm infection is regarded >10,000 EPG.

#### Exclusion criteria

Each patient was interviewed by qualified medical personnel. Patients who were screened as sero-positive for HIV were not part of this study. Patients with conditions that are well known to alter the haemoglobin level (pregnancy, sickle cell anaemia, etc.) were not included also. Patients on worm medicine, or haematinics, who have recently received blood transfusion, were excluded in this study.

### Data analysis

The results obtained was subjected to statistical analysis using SPSS package version 22. Descriptive statistics was used to calculate the prevalence and intensity of infections. Chi-square test was used to assess the associations and differences between the data on variables and the results from specimen examinations. Differences and associations was considered not significant at a probability (P) value greater than 0.001 (P > 0.001) and significant at a P value less than 0.001 (P < 0.001).

## RESULTS

A total of 200 subjects were recruited for this study, comprising 109 (54.5%) males and 91 (45.5%) females. Majority of the patients, 103 (51.5%) participants were within the ages of 31– 40 years. The age groups of 21-30 years and 41-50 years had 54(27%) and 43(21.5%) participants respectively (Table 1). The infection frequency was highest in the 31-40 years age group (24.3%) and lowest in the 41-50 years age group (22.2%). For the 21-30 years age group, the infection frequency was 22.2%. This, however, was not statistically significant (x^2^=0.217, p=0.897). It was also observed that double infections were more prevalent within the 21-30 years age group (11.1%) when compared to the 31-40 years age group (8.7%) and the 41-50 years age group (9.3%), whereas single infections were more prevalent within the 31-40 years age group (15.5%) than the 41-50 years age group (11.6%) and the 21-30 years age group (11.1%). Table 2 shows the prevalence of parasitic infection among different age groups.

**TABLE 1:** Age and sex distribution of patients studied.

| Age group | Anaemic group |  | Non-anaemic group |  | Total (%) |
| --- | --- | --- | --- | --- | --- |
|  | Male (%) | Female (%) | Male (%) | Female (%) |  |
| 21-30 years | 10(10.0) | 23(23.0) | 9(9.0) | 12(12.0) | 54 (27.0) |
| 31-40 years | 28(28.0) | 18(18.0) | 34(34.0) | 23(23.0) | 103 (51.5) |
| 41-50 years | 12(12.0) | 9(9.0) | 16(16.0) | 6(6.0) | 43 (21.5) |
| Total | 50(50.0) | 50(50.0) | 59(59.0) | 41(41.0) | 200 |

**TABLE 2:**
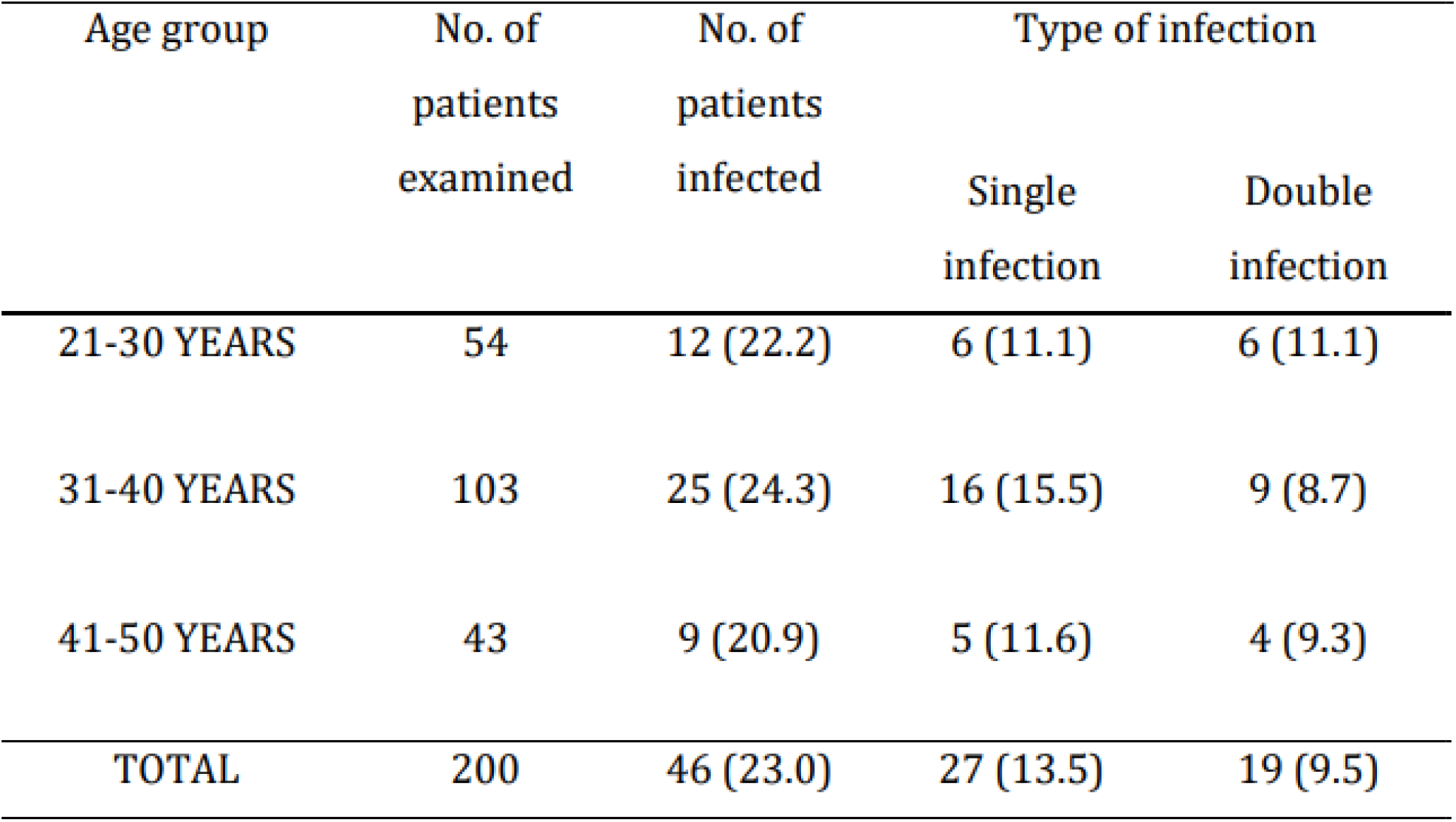
Prevalence of parasitic infection among different age groups.

| Age group | No. of patients examined | No. of patients infected | Type of infection |  |
| --- | --- | --- | --- | --- |
|  |  |  | Single infection | Double infection |
| 21-30 YEARS | 54 | 12 (22.2) | 6 (11.1) | 6 (11.1) |
| 31-40 YEARS | 103 | 25 (24.3) | 16 (15.5) | 9 (8.7) |
| 41-50 YEARS | 43 | 9 (20.9) | 5 (11.6) | 4 (9.3) |
| TOTAL | 200 | 46 (23.0) | 27 (13.5) | 19 (9.5) |

Five types of intestinal parasites were isolated from the stool samples of patients in this study viz *Entamoeba histolytica*, Hookworm species, *Ascaris lumbricoides, Trichuris trichiura*, and *Balantidium coli* (Table 3). Hookworm was the commonest intestinal parasite observed in the study with a total prevalence of 28 (14%), followed by *E. histolytica* 18 (9%), *A. lumbricoides* 10 (5%), *T. trichiura* 7 (3.5%), and *B. coli* 2 (1.0%). Prevalence of intestinal parasites according to gender revealed that males (26.6%) were more infected than females (18.7%) though this was not statistically significant (X^2^=1.758 p=0.185). Table 3 shows the prevalence of intestinal parasites according to gender.

**TABLE 3:** Prevalence of intestinal parasites according to gender.

| Gender | No. of patients examined | No. (%) of patients infected | No. (%) prevalence of the various intestinal parasites |  |  |  |  |  |
| --- | --- | --- | --- | --- | --- | --- | --- | --- |
|  |  |  | Hook worm | *E. histolytica* | *Trichuris trichiura* | *Ascaris lumbricoides* | *Balantidium coli* | Total |
| Male | 109 | 29 (26.6) | 17(15.6) | 12(11.01) | 7(6.42) | 4(3.67) | 1(0.92) | 41(37.61) |
| Female | 91 | 17 (18.7) | 11(12.09) | 6(6.60) | - | 6(6.60) | 1(0.10) | 24(26.37) |
| Total | 200 | 46 (23.0) | 28(14.00) | 18(9.00) | 7(3.50) | 10(5.00) | 2(1.00) | 65(32.50) |

Polyparasitism was observed in 19 (9.5%) of the cases consisting hookworm with *E. histolytica* 7 [males 5(4.59%) females 2(2.20%)], hookworm with *A. lumbricoides* 5 [males 2(1.83%) females 3(3.30%)], hookworm with *T. trichiura* 3 [males 3(2.75%)], *E. histolytica* with *T. trichiura* 2 [males 2(1.83%)], and *E. histolytica* with *A. lumbricoides* 2 [females 2(2.20%)](Table 4). The highest prevalence of polyparasitism was observed in males. All cases of Polyparasitism were anaemic (Table 4). The infection intensities of hookworm among patients were of moderate to high type (2000≥x<10000 epg) with one patient having a very high worm load (>10000 epg). Infections with *A. lumbricoides* and *T. trichiura* were observed to be more of low to moderate intensity (Table 5).

**TABLE 4:** Prevalence of polyparasitism in relation with gender of patients.

| Gender | no. of patients examined | NO. (%) OF POLYPARASITISM CASES |  |  |  |  |  |
| --- | --- | --- | --- | --- | --- | --- | --- |
|  |  | Hookworm +<br>*E. histolytica* | Hookworm +<br>*A. lumbricoides* | Hookworm +<br>*T. trichiura* | *E. histolytica* +<br>*T. trichiura* | *E. histolytica* +<br>*A. lumbricoides* | Total |
| Male | 109 | 5(4.59) | 2(1.83) | 3(2.75) | 2(1.83) | - | 12(11.01) |
| Female | 91 | 2(2.20) | 3(3.30) | - | - | 2(2.20) | 7(7.69) |
| Total | 200 | 7(3.50) | 5(2.50) | 3(1.50) | 2(1.00) | 2(1.00) | 19(9.50) |

**TABLE 5:**
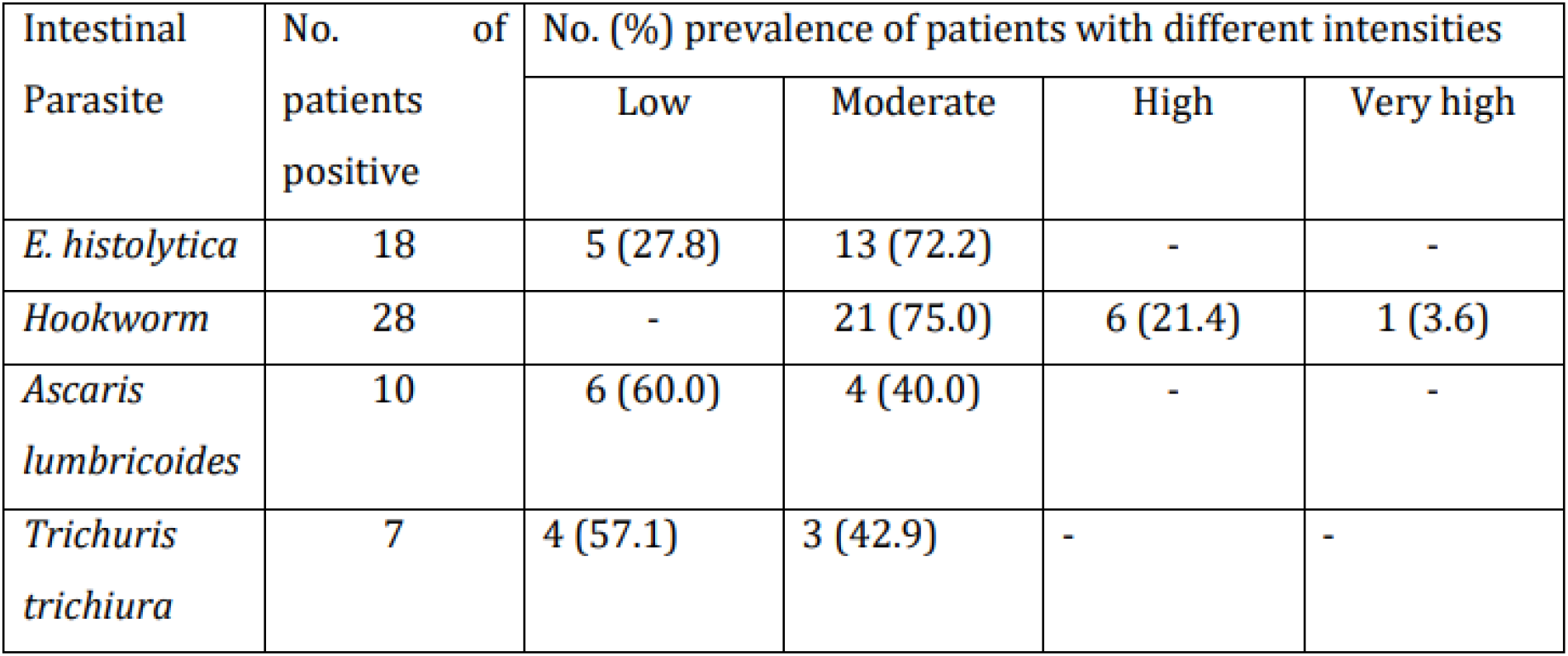
Intestinal parasite intensity based on egg counts (EPG)

| Intestinal Parasite | No. of patients positive | No. (%) prevalence of patients with different intensities |  |  |  |
| --- | --- | --- | --- | --- | --- |
|  |  | Low | Moderate | High | Very high |
| *E. histolytica* | 18 | 5 (27.8) | 13 (72.2) | - | - |
| *Hookworm* | 28 | - | 21 (75.0) | 6 (21.4) | 1 (3.6) |
| *Ascaris lumbricoides* | 10 | 6 (60.0) | 4 (40.0) | - | - |
| *Trichuris trichiura* | 7 | 4 (57.1) | 3 (42.9) | - | - |

The relationship between parasitic infection and haematological index (PCV %) is shown in Table 6. It showed an overall decrease in the mean PCV of individuals who were positive for any of the intestinal parasites found, when compared between the anaemic and non-anaemic groups. The PCV range of the patients was 11 to 50 % with a mean of 32.51%. Table 7 shows the mean PCV (%) of anaemic and non-anaemic patients examined in UCTH during the sampling period. There was a greater variation in the PCV values of the anaemic group. It ranged from 11% to 31% with a mean of 22.83%, while the PCV values of the non-anaemic group ranged from 36 % to 50% with a mean of 42.20% (Table 7).

**TABLE 6:** Age range of anaemic and non-anaemic patients in relation to the prevalence of intestinal parasites.

| Age group (years) | Status of patient | No. of patients examined | Mean PCV (%) | *E. histolytica* | Hookworm | *Ascaris lumbricoides* | *Trichuris trichiura* | *Balantidium coli* |
| --- | --- | --- | --- | --- | --- | --- | --- | --- |
| 21-30 | Anaemic | 33 | 22.76 | 9 (27.3) | 5 (15.2) | 3 (9.1) | 0 (0.0) | 0 (0.0) |
|  | Nonanaemic | 21 | 41.19 | 0 (0.0) | 1 (4.8) | 0 (0.0) | 0 (0.0) | 0 (0.0) |
|  | Total | 54 | 29.93 | 9 (16.7) | 6 (11.1) | 3 (5.6) | 0 (0.0) | 0(0.0) |
| 31-40 | Anaemic | 46 | 22.33 | 5 (10.9) | 15 (32.6) | 3 (6.5) | 7 (15.2) | 1 (2.2) |
|  | Nonanaemic | 57 | 42.40 | 1 (1.8) | 2 (3.5) | 0 (0.0) | 0 (0.0) | 0 (0.0) |
|  | Total | 103 | 33.44 | 6 (5.8) | 17 (16.5) | 3 (2.9) | 7 (6.8) | 1 (1.0) |
| 41-50 | Anaemic | 21 | 24.05 | 3 (14.3) | 5 (23.8) | 4 (19.4) | 0 (0.0) | 1 (4.8) |
|  | Nonanaemic | 22 | 42.64 | 0 (0.0) | 0 (0.0) | 0 (0.0) | 0 (0.0) | 0 (0.0) |
|  | Total | 43 | 33.56 | 3 (7.0) | 5 (11.6) | 4 (9.3) | 0 (0.0) | 1 (2.3) |
| TOTAL | Anaemic | 100 | 22.83 | 17 (17.0) | 25 (25.0) | 10 (10.0) | 7 (7.0) | 2 (2.0) |
|  | Nonanaemic | 100 | 42.20 | 1 (1.0) | 3 (3.0) | 0 (0.0) | 0 (0.0) | 0 (0.0) |
|  | Total | 200 | 32.51 | 18(9.0) | 28 (14.0) | 10 (5.0) | 7 (3.5) | 2 (1.0) |

**TABLE 7:** Mean PCV (%) of anaemic and non-anaemic patients examined in UCTH during the sampling period.

| Status of patients | No. of patients examined | Mean PCV (%) | Standard deviation | Maximum PCV (%) | Minimum PCV (%) | Range | Variance | % of total |
| --- | --- | --- | --- | --- | --- | --- | --- | --- |
| Anaemic | 100 | 22.83 | 4.878 | 31 | 11 | 20 | 23.799 | 50.0% |
| Nonanaemic | 100 | 42.20 | 3.247 | 50 | 36 | 14 | 10.545 | 50.0% |
| Total | 200 | 32.51 | 10.553 | 50 | 11 | 39 | 111.357 | 100.0% |

Forty-two (42) patients (42.0%) were generally infected within the anaemic group. Single infections i.e. infections caused by only one intestinal parasite was observed in 23 (23.0%) patients while double infections (caused by two coexisting intestinal parasites) were observed in 19 (19.0%) patients within the anaemic group. In the non-anaemic group only 4 (4.0%) patients were infected with intestinal parasites and all 4 infections were single infections. The prevalence of intestinal parasite infection between the anaemic and non-anaemic group was very highly significant (x^2^=40.768, p<0.00001) (Table 8).

**TABLE 8:** Prevalence of parasitic infection between anaemic and non-anaemic patients.

| Status of patient | No. of patients examined | No. of patients infected | Type of infection |  |
| --- | --- | --- | --- | --- |
|  |  |  | Single infection | Double infection |
| Anaemic | 100 | 42 (42.0) | 23 (23.0) | 19 (19.0) |
| Non-anaemic | 100 | 4 (4.0) | 4 (4.0) | 0 (0.0) |
| Total | 200 | 46 (23.0) | 27 (13.5) | 19 (9.5) |

## DISCUSSION

The overall prevalence of intestinal parasites in this study was found to be 23% (n=200). This was lower than that observed among pre-school children in Calabar (49.7%) [15]. This difference in prevalence rates was expected since the subjects in this study were all adults ranging from 21 to 50 years of age. It is well established that intestinal parasites can affect all age groups and gender, but the situation is more prevalent among toddlers and children. This is widely due to their lower immune response compared to adults, poor hygiene, and poor sanitary and environmental conditions which favour the development of parasites and eventual infection of hosts [16]

There was no significant difference in intestinal parasite infection of the different age groups but the highest prevalence (24.3%) was found among the age group of 31-40 years. This disagrees with [17], who reported that incidence rates decreased progressively with age (less in 31-50years). Prevalence rates reaching 49.7% among children aged 1-5 years in Calabar [15]; 92.8% among 1-4 years, 69% among 5-9 years, 34.6% among 10-15 years in Ibadan [18]; 60.7% among 1-5 years, 25.9% among 6-10 years and 13.4% among 11-15 years in Edo state [19] have been reported. Studies done in different parts of the world also revealed high prevalence of infection among children [20][21]. The susceptibility of children to parasitic infections is due to their lower immune response compared to adults, poor hygiene, and poor sanitary and environmental conditions which favour the development of parasites and eventual infection of hosts [16].

Higher parasitic infections were observed among males (26.6%) compared to females (18.7%) in this study although the difference was not statistically significant. This gender relationship with parasite infection agrees with [15], but disagrees with [17] and [19]. This pattern of parasitism with respect to gender could best be attributed to a higher level of hygiene normally observed among the females of this age bracket compared to their male counterpart. Moreover, the higher prevalence rate observed in males in this study reflects the greater exposure of males to infection. Males are generally more active, less careful and less discriminatory about what they eat which might make the spread of the infection easier and faster. Another reason could be attributed to factors such as buying of food from vendors who may have poor hygiene and consumption of inadequately cooked vegetables.

Hookworm was the commonest parasite infecting patients in our study (14%) followed by *E. histolytica* (9%), *Ascaris lumbricoides* (5%), and *Trichuris trichiuria* (3.5%). *Balantidium coli* and *Diphyllobothrium latum* infections were each observed in 1 out of the 200 patients (0.50%). Overall, the infection rates of parasites were lower than those obtained in other reports [15][22], especially those working on a younger population. The presence of the common triad *A. lumbricoides, T. trichiur*a and hookworm is comparable with many previous reports in Nigeria [23][24][25][26][27][28][15][19]. These parasites did have a toll on patients. The mean PCV (%) of infected and uninfected patients in this study were observed to be 24.78 and 34.82 respectively. This could be explained by the methods in which the various parasites caused anemia. It is reported that an adult hookworm ingest about 0.03ml to 0.2ml of blood daily [29]. The prevalence rate of hookworm in this study (14%) matches that in Abeokuta (13.9%) [25]. It is slightly lower than that reported by [19], who recorded 16.90% prevalence among children in rural communities of Edo State, Nigeria, and [30], who had 17%, but higher than that reported by [15] in Calabar, Nigeria. Higher prevalence rates of hookworm infection (43.75%) have been reported among adolescent boys [22], 45% in a Lagos suburb [31].

In this study, hookworm infection was the only infection observed to range from moderate intensity to very high intensity concerted by egg counts of 2000 to 10000 epg). Three potential factors remain as significant predictors of hookworm infection: farming, lack of a closed latrine, and zone of residence [32]. Although these factors were not investigated as part of this study, it is worth mentioning that an important determinant of hookworm infection is working as a farmer. Farmers often work with bare feet in the field, making them vulnerable to penetration by infective hookworm larvae. The use of untreated human faeces in agricultural fields concentrates hookworm eggs in a location where women may be exposed on a daily basis while tending their farms, causing hookworm prevalence to peak in adulthood [33].

Hookworm is an important cause of anaemia worldwide [34]. A high significant difference was observed in hookworm infection between the anaemic group and the non-anaemic group (x^2^=20.1, p<0.001). [35] reported a significant negative correlation between plasma ferritin levels and hookworm load. Even a low hookworm load can cause anaemia in people whose intake of iron is low and whose iron stores are already depleted [36].

The different incident rates of hookworm infection between males and females were not significant statistically (x2=0.507, p=0.476). Nevertheless, infections were slightly higher in males than females (15.6% and 12.1% respectively). This trend is consistent with the findings of [37] but contradicts that of [17]. The age group of 31-40 years had the highest infection (16.5%) compared to the other age groups (11.6% for 41-50 years and 11.1 % for 21-30 years). This however, was not statistically significant (x^2^=1.112, p = 0.573). Hookworm prevalence and intensity can be higher among adults since the infection tends to be occupational, making farmers, plantation workers and coal miners more susceptible [38].

*Trichuris trichiura* was present at an overall prevalence of 3.5%. All infections occurred within the anaemic group [7% (n=100)]. Infections were more of low intensity than moderate intensity (4% and 3% respectively). Infections were observed only among the male counterparts and only within the age group of 31-40 years. The low prevalence of 3.5% in this study is incompatible with those of [30], and [19], but is congruent with the report of [39], who not only observed *T. trichiura* infection to be the lowest among other parasites in Ijebu-Ode but also observed that all patients who were infected with it were anaemic. High intensity *Trichuris* infection is known to influence nutritional status [34] but [40] also advocated that *Trichuris* is related to anaemia not only in severe infection but also in mild infection. *Trichuris* is suggested to be associated with anaemia mediated through iron deficiency caused by blood loss or anorexia [11]. [17] reported the incidence rate to be higher among the age groups 10-20 and 21-30 respectively, and decreased progressively with age (less in 31-50 and 51-65 age groups), being higher in females than in males. The decrease in the prevalence of *T. trichiura* among the older age group in this study may reflect a change in the hygiene behaviour. It was also observed in this study that only 2(28.6%) *Trichuris* infections occurred as single infections. The majority were joint infections with either hookworm 3(42.9%) or *E. histolytica* 2(28.6%). *Trichuris* was also reported in much lower frequencies, often in combination with other helminths [41]. *T. trichiura* is more likely to be concurrent than expected by chance [32]. [42] reported that children with concomitant *T. trichiura* and hookworm infections had lower blood haemoglobin levels than children with neither or only one of these parasites. Co-infection with hookworm or *E. histolytica* seems to aggravate trichuriasis symptoms. It often occurs along with hookworm infections and so may accelerate the onset of iron-deficiency anaemia [11][12]. The mean PCV of those infected with *Trichuris*, and those not infected in this study were observed to be 23.86% and 32.86 % respectively.

There was a significant association between infection with *Ascaris lumbricoides* and anaemia. The mean PCV of those with ascariasis and those not infected were observed to be 23.80% and 32.97% respectively. The prevalence of 5% for *Ascaris* in this study confirms the effects of the worm to a minority who may still lack good sanitation and adequate hygiene habits. In prior reports, the prevalence of Ascariasis has gone as high as 75%, where a high occurrence of unhygienic habits exist [30][19]. *A. lumbricoides* eggs are very resistant to harsh environmental conditions and are airborne. They may account for the ubiquitous nature of egg distributions and hence very high prevalence in all the age groups. A higher number of females 6 (6.6%) than males 4 (3.7%) were observed to be infected in this study. Infection prevalence rates were higher in the age group of 41-50 years (9.3%) and 21-30 years (5.6%) and least in the age group of 31-40 years. The prevalence of *Ascaris* infection in this study increased with age. The infection intensity for this study was observed to be mild. Ascariasis was significantly associated with anaemia (x^2^=10.526, p=0.001) though it was concurrent with either hookworm or *E. histolytica* infections. *Ascaris lumbricoides* infections are commonly asymptomatic, although clinical complications of extra-intestinal or high numbers of *Ascaris* may accelerate the onset of iron deficiency [11][12].[19] also reported a significant association between *Ascaris lumbricoides* and hookworm infection and anaemia (*P* < .001).

*E. histolytica* infection with an overall prevalence of 9% was the second most prevalent disease condition observed in this study. The infection was highly significantly associated with the anaemic group (p<0.001). The younger age group, 2130 years had the highest incident rate (16.7%) as it then decreased with age. This was similarly observed by [17]. Again infection was higher in males (11%) than females (6.6%) though this was not statistically significant. Faecal contaminated water, food or hands or via anal sexual contact are possible transmission routes. Polyparasitism between *E. histolytica* and hookworm was higher (38.9%) than with *Trichuris trichiura* (11.1%). A prevalence rate of 24% has been reported among the age groups of 10-20 years and 21-30 years in a rural setting [17].

Least common infections in this study were those of *Balantidium coli* and *Diphyllobothrium latum*. Both were observed in 1 out of 200 (0.50%). Though not enough to draw conclusion on any significant association with anaemia, the presence of *B. coli* suggests a possible contamination of water source.

This study has confirmed the existence of a relationship between intestinal parasite infections and alterations in haematological indices resulting in anaemia. A greater number of the anaemic patients [42% (n=100)] in the present study were infected with one or more intestinal parasites, whereas only [4% (n=100)] were infected in the non-anaemic group. This is in accordance with an earlier report by [17] who presented proof of parasitic infections having adverse effect on haemoglobin, packed cell volume, and indirectly on nutritional status. This relationship between intestinal parasite infection and anaemia has reflected in many reports from different regions [15][21][19].

It was also observed that more than half of the anaemic subjects (58%) did not have intestinal parasites, which suggests the presence of other factors that could cause anaemia other than parasitic infections. Possible factors may be: dietary iron deficiency; deficiencies of other key micronutrient including folate, vitamin B_12_ and vitamin A, riboflavin, and copper; inflammatory disorders, haemorrhage, lactation; drugs: e.g. dapsone; infectious diseases such as malaria, schistosomiasis, cancer, tuberculosis; or inherited conditions that affect red blood cells (RBCs), such as thalasemia, [4].

There was a significant difference between the mean PCV if infected patients and uninfected patients (p<0.001). Insufficient intake of iron in diet results in lower levels of hemoglobin as it contains iron. Iron deficiency anaemia in itself could be not only by insufficient dietary intake of iron, but also by gastro-intestinal tract bleeding due to parasitic infections and malabsorption of nutrients in the intestines [3]. Although, not all cases of parasite infection resulted in anaemic conditions as observed in the nonanaemic group who had a prevalence of 4% for intestinal parasites.

## CONCLUSION

This study has demonstrated that intestinal infections and anaemia are common health problems. Attention is hereby needed to tackle this avoidable misery and reduce significantly their toll on mankind. There are many affordable and costeffective strategies to improve the situation. They include education, deworming, improving hygiene, water and sanitation, and nutrient supplements. However, these efforts should be accompanied by the long term improvement in the under lying determinants of undernutrition, such as poverty and lack of awareness. However, poor sanitation, under-nutrition, inadequate personal and domestic hygiene contribute immensely to the none-effectiveness of educational standard in reducing positive rates of soil transmitted helminth (STH) infections. Since this study was done on a population limited to those patients attending the University of Calabar teaching hospital, it does not claim to have assessed the anaemia prevalence among the entire populace. However, it does point the fact that anaemia is prevalent in the region equally affecting males and females of all ages.

WHO guidelines for the evaluation of parasitic infection at the community level only recommend mass treatment for primary and secondary school children [43]. It is therefore recommended that health education on major tropical diseases should not be limited to rural areas or primary and post primary institution only as it is usually practiced. Adequate focus should be given to improve sanitation and excreta disposal system to control the spread of intestinal parasites in our environment.

## Data Availability

All data produced in the present work are contained in the manuscript

## ACKNOWLEDGEMENT

The authors are grateful to the technical staff of the University of Calabar Teaching Hospital for their support during the study.

